# Long-Term Subarachnoid Hemorrhage Risk and Survival after Microsurgical Clipping of Unruptured Intracranial Aneurysms

**DOI:** 10.1101/2025.09.23.25336517

**Authors:** Shunsuke Kawamoto, Go Ikeda, Shunsuke Fukaya, Kanae Okunuki, Hiroyoshi Akutsu

## Abstract

**Background and Purpose:** Prophylactic microsurgical clipping for unruptured intracranial aneurysms (UIAs) effectively prevents rupture, yet its long-term impact on cerebrovascular and systemic health remains uncertain. We assessed subarachnoid hemorrhage (SAH) risk, cerebrovascular outcomes, and systemic disease incidence after clipping under stringent surgical selection criteria.

**Methods:** We retrospectively analyzed 930 patients (990 procedures) with asymptomatic anterior-circulation UIAs treated between 2003–2025, totaling 7,638 patient-years (median follow-up 8.3 years). The primary endpoint was postoperative SAH; secondary endpoints included all-stroke, malignancy, cardiovascular disease, dementia, and all-cause mortality. Incidence rates (per 1,000 patient-years) with 95% CIs were derived using Byar’s approximation; rate ratios (RRs) versus natural-history cohorts (UCAS, SUAVe) and standardized incidence ratios (SIRs) versus Japanese population registries were computed using exact Poisson methods.

**Results:** Ten SAH events occurred (1.31/1,000 patient-years; 95% CI, 0.63–2.41). Compared with UCAS and SUAVe, clipping reduced SAH risk by 86–93% (RR 0.14 vs UCAS; 0.24 vs SUAVe; both p<0.001), with the lowest risk in patients without untreated aneurysms (0.64/1,000 patient-years). Nevertheless, age-adjusted analyses revealed persistent cerebrovascular vulnerability: SAH incidence exceeded the general population (SIR 3.22; 95% CI, 0.88–8.25; p=0.075), and all-stroke incidence was doubled (SIR 2.11; 95% CI, 1.50–2.90; p<0.001). Dementia incidence in the ≥70-year subset was significantly lower (SIR 0.34; 95% CI, 0.20–0.53; p<0.001). Cardiovascular disease risk was unchanged (SIR 1.41; p=0.221). A modest malignancy excess (SIR 1.36; p=0.009) likely reflected surveillance bias rather than treatment-related risk.

**Conclusions:** Microsurgical clipping provides durable long-term protection against SAH, reducing rupture risk by >90% versus natural history. However, residual cerebrovascular risk—driven partly by de novo aneurysm formation—underscores the need for lifelong vascular risk management and imaging surveillance even after successful aneurysm treatment.

## INTRODUCTION

The primary objective of prophylactic treatment for unruptured intracranial aneurysms (UIAs) is to prevent future subarachnoid hemorrhage (SAH). Direct microsurgical clipping has long been performed as an established treatment method, but its effectiveness has traditionally been evaluated mainly in terms of angiographic occlusion rates and perioperative complications, with limited comprehensive assessment of long-term health outcomes.

Large-scale natural history studies, such as the Unruptured Cerebral Aneurysm Study (UCAS)^1^ and the Small Unruptured Intracranial Aneurysm Verification Study (SUAVe)^2^, have provided valuable insights into the rupture risk of UIAs, generally reporting annual rupture rates of less than 1%, particularly low for small lesions. Consequently, current guidelines recommend conservative observation for many small aneurysms^3^. Nevertheless, harboring an aneurysm itself can represent a substantial psychological burden for patients^4^.

From another perspective, patients with UIAs frequently present with vascular risk factors such as hypertension, smoking, and family history, which not only contribute to aneurysm formation^5,6^ but may also predispose them to cerebrovascular diseases in general, even after treatment. At the same time, regular medical observation may facilitate earlier detection and treatment of other diseases, making the overall impact on long-term health status more complex.

Despite advances in endovascular techniques, the optimal management strategy for UIAs remains controversial, particularly for small lesions in younger patients^7,8^. Previous surgical series have predominantly focused on perioperative or angiographic outcomes, with limited attention to broader, long-term health impacts including neuropsychological or psychosocial outcomes^9^. Moreover, most studies include heterogeneous patient populations varying significantly in operative risk, complicating the assessment of surgical effectiveness under ideal conditions^10^.

To address this knowledge gap, we conducted a retrospective study focusing specifically on highly selected patients with anterior circulation UIAs judged to carry minimal operative risk. By restricting the cohort to cases most suitable for clipping, we aimed to assess the long-term effectiveness and safety of surgical intervention under ideal circumstances, while acknowledging the inherent limitations in generalizability.

## MATERIALS AND METHODS

### Study Population and Setting

This study was conducted and reported in accordance with the STROBE (Strengthening the Reporting of Observational Studies in Epidemiology) guidelines for observational cohort studies. A completed STROBE checklist with manuscript line numbers is provided in the Supplemental Material^11^. This retrospective observational study included patients who underwent microsurgical clipping for asymptomatic UIAs of the anterior circulation between January 2003 and July 2025. All cases were treated at Dokkyo Medical University from January 2003 to March 2023 and subsequently at Nasu Red Cross Hospital after the first author’s relocation in April 2023. Identical surgical indications, operative techniques, and follow-up protocols were maintained across both institutions, thereby providing a single continuous cohort under a unified methodology.

### Patient Selection Criteria and Study Cohort

Patients were included if they had anterior-circulation UIAs treated by microsurgical clipping between April 2003 and July 2025 with available postoperative imaging and at least 5 years of follow-up. Detailed surgical indications and technical protocols have been described previously^12^ and are only summarized here. Patients with posterior-circulation aneurysms, symptomatic aneurysms, giant aneurysms ≥25 mm, residual domes requiring early additional treatment, or insufficient clinical records were excluded.

During the study period, a total of 1,040 microsurgical clipping procedures for anterior-circulation UIAs were performed in 979 patients. Five patients were excluded: two patients with previously ruptured aneurysms treated by endovascular means, who also had concomitant unruptured aneurysms clipped after recovery from SAH; however, the originally ruptured lesions re-ruptured during follow-up (IC-PC and BA top, one each), leading to death, and three with early postoperative neck remnants (Acom 1, IC-PC 1, ICA C2 1) requiring additional endovascular treatment during the early postoperative period. After exclusion of these five cases, 1035 procedures in 974 patients remained eligible. Subsequently, 33 patients with remote follow-up and 11 with early postoperative dropout were excluded, leaving a final source population of 930 patients undergoing 990 procedures (655 females, 275 males; mean age, 62.8 ± 10.5 years; range, 27–86). Because some patients underwent separate operations for aneurysms treated at different times and some procedures addressed multiple aneurysms simultaneously, the total number of procedures exceeded the number of patients, and the total number of aneurysms exceeded the number of procedures.

### Surgical Team, Surgical Technique and Intraoperative Safety Strategies

Nearly all procedures were performed by the first author (S.K.), a senior cerebrovascular neurosurgeon with extensive experience (>500 prior clipping cases). Other staff neurosurgeons with >100 prior cases served as primary operators under S.K.’s direct supervision.

All procedures were performed using a standardized microsurgical protocol as previously described. Briefly, a clipping-first approach was employed for anterior-circulation aneurysms, with surgical strategy and intraoperative monitoring (including MEP) applied according to established institutional protocols^13–19^. Brain-protective measures, temporary proximal control, and indocyanine green videoangiography were routinely used. When anatomical complexity or perforator involvement posed a risk to neurological function, a safety-first approach allowing intentional preservation of small neck remnants was adopted to minimize morbidity.

### Comprehensive Follow-up Protocol

All patients were routinely followed in the outpatient clinic under direct care of the first author, ensuring continuity and uniformity of clinical and radiological assessments. Standardized follow-up intervals were scheduled at 1, 3, 6, and 12 months postoperatively, and annually thereafter. Each visit included structured neurological examination, MRI/MRA evaluation, systematic cardiovascular risk factor assessment, and review for new neurological symptoms.

Study-specific surveillance: Long-term imaging surveillance consisted of annual MRI/MRA for the first 10 years, followed by biennial imaging. CTA or DSA was scheduled every 5 years or earlier if clinically indicated. Additional imaging was obtained when recurrence, de novo aneurysm formation, or enlargement of untreated aneurysms was suspected.

### Data Collection and Event Definition

Information on newly diagnosed systemic or neurological diseases was prospectively captured through the electronic medical record system. Disease definitions were: stroke (cerebral infarction or intracerebral hemorrhage confirmed by neuroimaging); cardiovascular disease (ischemic heart disease or heart failure requiring hospitalization); malignant neoplasms (newly diagnosed cancers confirmed pathologically); and dementia (clinical diagnosis by neurologists or psychiatrists based on standardized criteria).

### Outcome Measures

The primary outcome was postoperative SAH. Secondary outcomes included all-stroke occurrence, malignant neoplasm, cardiovascular disease, dementia, and all-cause mortality. Follow-up duration was defined from surgery date to last follow-up or death. Patients who discontinued follow-up were handled as censored at the date of last clinical contact. Cases with documented reasons for discontinuation (e.g., advanced age, death from unrelated causes, relocation, treatment for other comorbidities) were treated as censored observations. True loss-to-follow-up cases, defined as patients with no subsequent clinical information, were included in sensitivity analyses to evaluate the potential impact of attrition bias on late outcomes.

### Statistical Analysis

Statistical analyses were performed using SPSS Statistics version 30.0.0.0 (IBM Corp., Armonk, NY, USA) and Python version 3.11 (Python Software Foundation). Annual incidence rates for each endpoint—SAH, all-stroke (including cerebral infarction and intracerebral hemorrhage), malignant neoplasms, cardiovascular disease, and dementia—were calculated as the number of events divided by the total observation period (person-years). Ninety-five percent confidence intervals (95% CIs) for single-group rates were derived using Byar’s approximation.

#### Population comparisons

For comparisons with the general population, we used representative Japanese registries to obtain reference rates harmonized to per 1,000 person-years: Cancer Statistics in Japan 2021 for all cancers and malignant brain/CNS tumors^20^, CIRCS for ischemic heart disease^21^, the Hisayama Study for dementia^22^, and BTRJ for benign primary brain tumors^23^. When age bands in the reference data did not exactly match our cohort, the closest available strata or weighted averaging were applied. For dementia, analyses were restricted to the ≥70-year subset in our cohort to align with Hisayama (≥65 years). Standardized incidence ratios (SIRs) were calculated as observed events / expected events, where expected events = reference rate × person-years / 1,000; two-sided exact Poisson tests provided p-values, and 95% CIs for SIRs used Byar’s approximation. Where contemporaneous age-specific population rates were unavailable, only crude (unadjusted) odds ratios (ORs) with Wald 95% CIs were reported^20,21^.

#### Cohort and cross-cohort comparisons

Rate ratios (RRs) between groups were computed under Poisson assumptions, with Wald 95% CIs on the logarithmic scale. For age-adjusted analyses of SAH and all-stroke, SIRs were additionally calculated using age-stratified incidence rates from Tochigi Prefecture^24^.

For cross-cohort comparisons, expected SAH events were estimated from the overall annual rupture rates reported by UCAS Japan (0.95% per year)^1^ and SUAVe (0.54% per year)^2^ and applied to our observed person-years (expected = rate × person-years / 1,000). Observed events divided by these expected counts yielded SIRs with 95% CIs by Byar’s approximation. Because detailed age-specific rupture rates were unavailable in UCAS and SUAVe, this indirect standardization partially accounted for age distribution but did not allow strict direct comparisons.

#### Time-to-event analyses

Time-to-event outcomes were analyzed using the Kaplan–Meier method, with survival probabilities and 95% CIs calculated using Greenwood’s formula. Group differences were assessed with the log-rank test, and SAH-free survival at 5 and 10 years was reported with numbers at risk at prespecified intervals.

#### Sensitivity analyses

Given the small number of SAH events, we performed several sensitivity analyses: bootstrap resampling (10,000 iterations) to confirm stability of incidence rate estimates; Bayesian modeling incorporating informative priors from UCAS^1^ and SUAVe^2^ to estimate the posterior probability of reduced SAH risk after clipping; statistical power analysis confirming 80% power to detect a 50% reduction in SAH incidence compared with UCAS^1^; and calculation of the minimum detectable effect size for age-adjusted comparisons with the general population, including the borderline finding (p = 0.075) ^24^. Because these analyses were exploratory, no multiplicity adjustment was applied; all p-values are presented descriptively, with two-sided p<0.05 considered statistically significant.

### Ethical Approval

This study was conducted in accordance with the Declaration of Helsinki and approved by the Institutional Review Boards of both participating institutions: Dokkyo Medical University (Approval No. R-81-2J) and Nasu Red Cross Hospital (Approval No. 2025-04). Written informed consent was obtained from all patients.

## RESULTS

### Patient Characteristics

Table 1 shows the background factors of the 930 patients analyzed. The mean age was 62.8±10.5 years, with women accounting for 70.4%. SAH family history was found in 309 cases (33.2%), personal SAH history in 43 cases (4.6%), hypertension in 571 cases (61.4%), and smoking history in 183 cases (19.7%). Multiple aneurysms were found in 273 cases (29.4%).

**Table 1.**
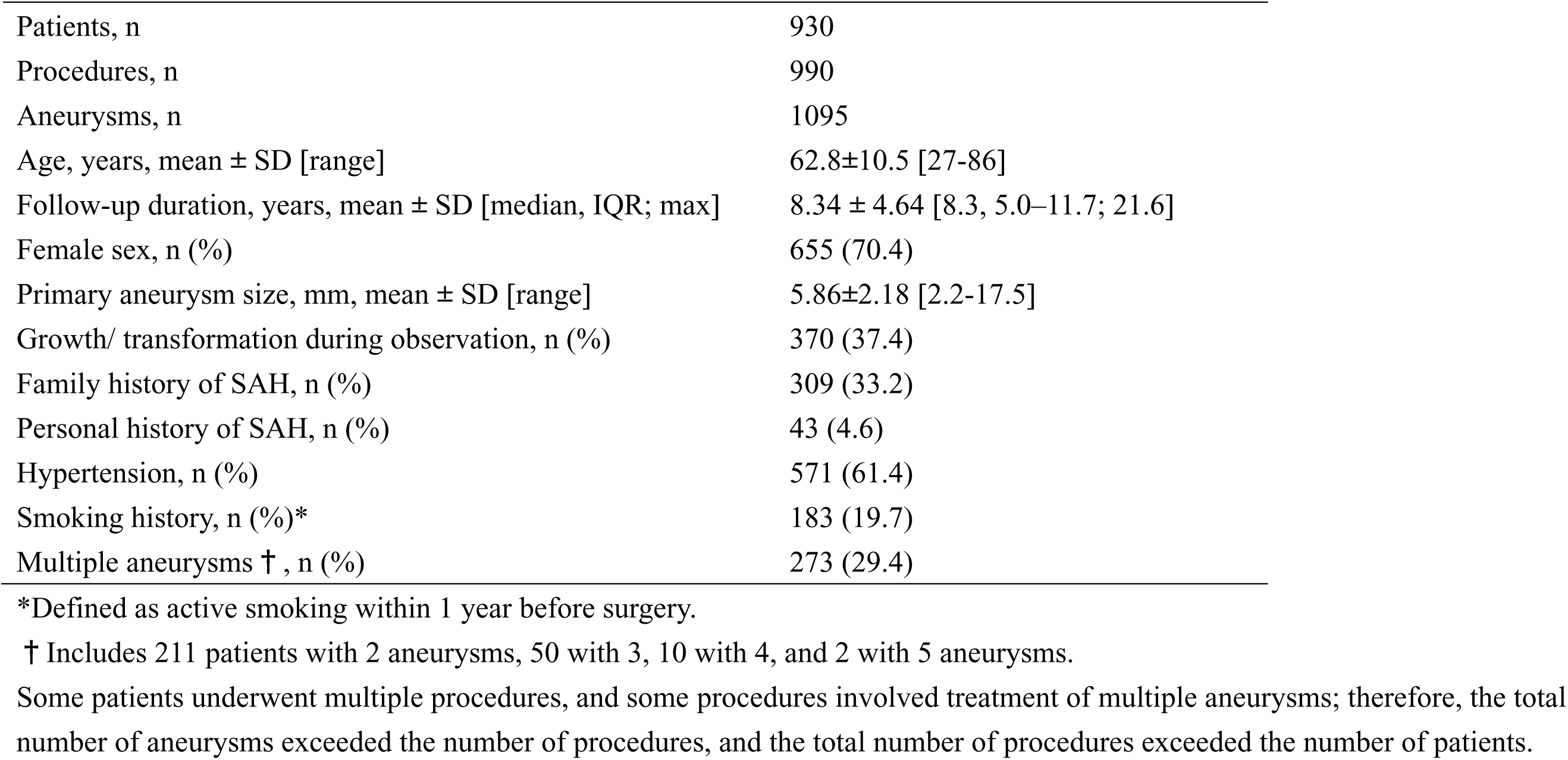
Patient characteristics.

Surgery was performed in 990 operations, with 1 aneurysm treated per operation in 893 cases (90.2%), 2 aneurysms in 89 cases (9.0%), and 3 aneurysms in 8 cases (0.8%). Cases that became surgical indications due to enlargement during observation numbered 370 cases (37.4%).

### Follow-up Status

The total observation period was 7,638 patient-years (PY). The median follow-up duration was 8.3 years (IQR, 5.0–11.7; maximum, 21.6 years), with a mean of 8.34 ± 4.64 years. Scheduled follow-up was completed in 675 patients (72.6%), whereas 255 patients (27.4%) discontinued follow-up during the study period. Among these, 100 patients (10.7%) were classified as lost to follow-up with no subsequent clinical information available, while 155 patients (16.7%) were censored because of advanced age (n = 33), death from unrelated causes (n = 39), relocation to distant regions (n = 20), or prioritization of treatment for other comorbidities (n = 63). Censored cases were treated as end-of-follow-up in all survival analyses.

### SAH Incidence and Survival Analysis

Immediate postoperative outcomes have been reported elsewhere; briefly, there was no perioperative mortality, and transient morbidity occurred in 1.5% of patients, all of whom recovered to functional independence (mRS 0–1) within 3 months. During the follow-up period, 10 SAH events occurred (Table 2), corresponding to an annual incidence rate of 1.31 per 1,000 patient-years (95% CI, 0.63–2.41; Byar). Of these, 5 cases (50.0%) resulted from rupture of untreated aneurysms, 4 (40.0%) from rupture of de novo aneurysms, and 1 (10.0%) from local recurrence at a previously treated site, defined consistently with the companion local-recurrence analysis. Notably, in Case 1, local recurrence occurred 9 years after clipping surgery for an ICPC aneurysm; coil embolization was subsequently performed, but further recurrence developed 4 years later, ultimately resulting in rupture. Detailed incidence rate comparisons with natural-history cohorts and the general population are summarized in Table 3.

**Table 2.** SAH Events During Follow-up.

| Case | Location | Type | Time to event (months) | Outcome |
| --- | --- | --- | --- | --- |
| 1 | Same location | Local recurrence* | 163 | mRS 1 |
| 2 | Same-side ICPC | Untreated aneurysm | 61 | mRS 5 |
| 3 | ACoA | De novo | 44 | mRS 0 |
| 4 | Left A2-3 | Untreated aneurysm | 129 | Death |
| 5 | Contralateral ICPC | De novo | 100 | mRS 0 |
| 6 | ICPC | De novo | 82 | mRS 1 |
| 7 | Contralateral ICPC | Untreated aneurysm | 37 | mRS 2 |
| 8 | BA top | De novo | 99 | mRS 1 |
| 9 | ICPC | Untreated aneurysm | 3 | mRS 0 |
| 10 | IC-OphA | Untreated aneurysm | 2 | Death |

Local recurrence refers to any aneurysmal regrowth at a previously treated site, defined as in the companion analysis of long-term recurrence risk after clipping (see Methods). ICPC = internal carotid–posterior communicating artery; ACoA = anterior communicating artery; BA = basilar artery; OphA = ophthalmic artery; mRS = modified Rankin Scale.

**Table 3.**
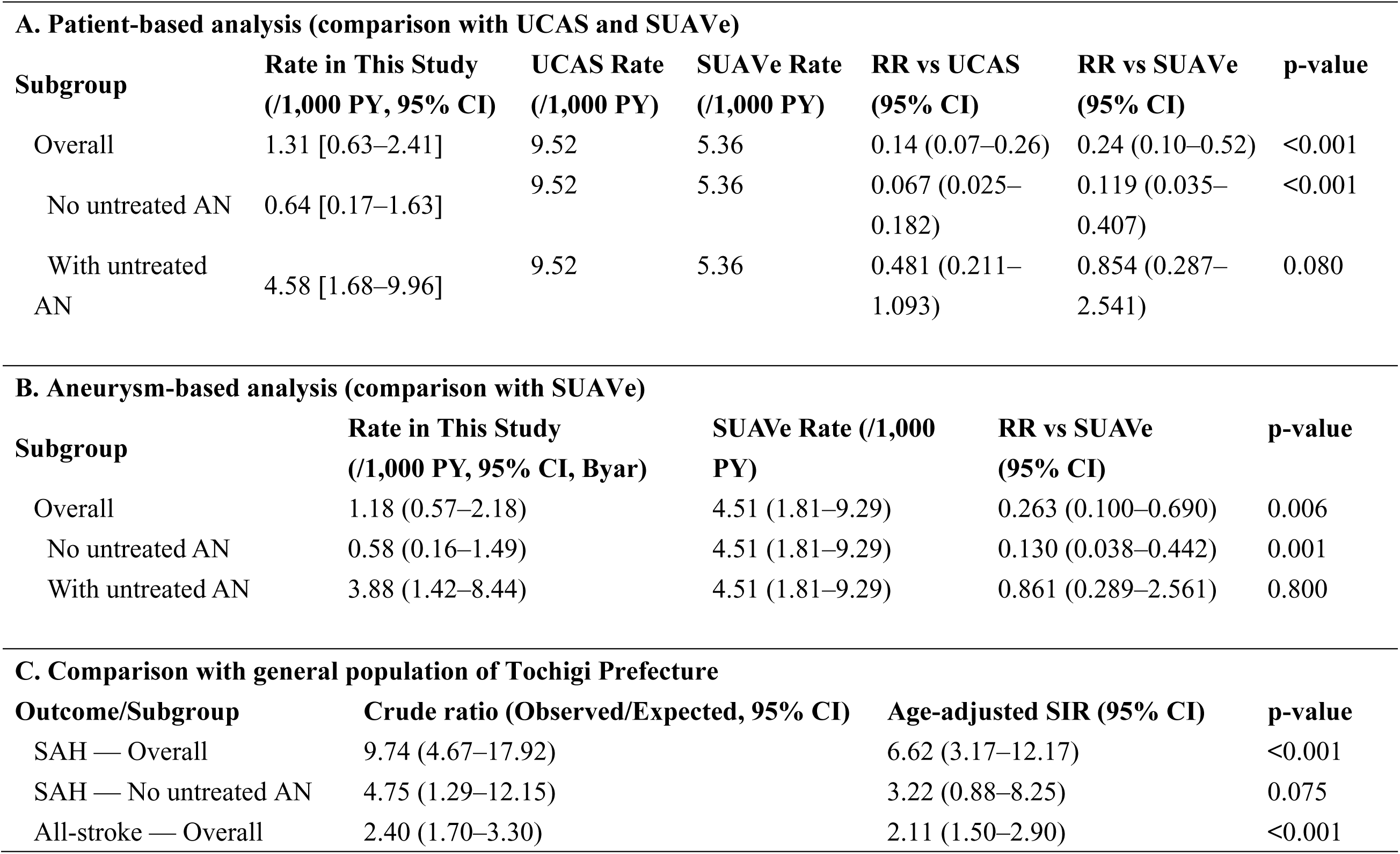
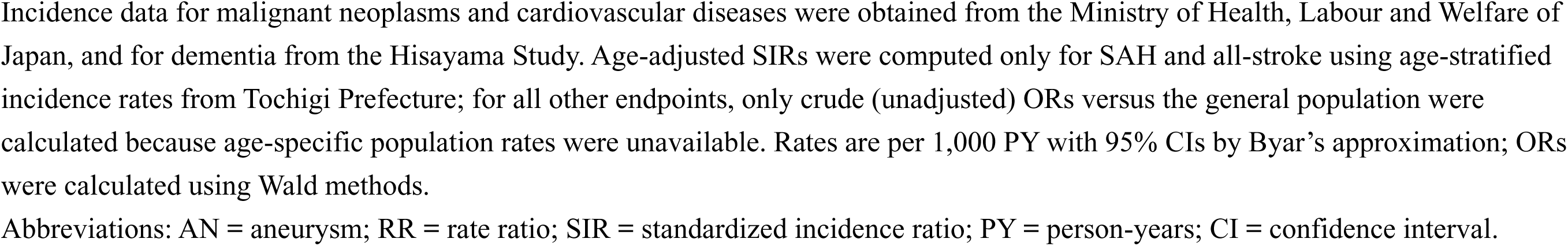
Comparison of SAH and Stroke Incidence Rate (Byar CIs)

| A. Patient-based analysis (comparison with UCAS and SUAve) |  |  |  |  |  |  |
| --- | --- | --- | --- | --- | --- | --- |
| Subgroup | Rate in This Study<br>(/1,000 PY, 95% CI) | UCAS Rate<br>(/1,000 PY) | SUAve Rate<br>(/1,000 PY) | RR vs UCAS<br>(95% CI) | RR vs SUAve<br>(95% CI) | p-value |
| Overall | 1.31 [0.63–2.41] | 9.52 | 5.36 | 0.14 (0.07–0.26) | 0.24 (0.10–0.52) | <0.001 |
| No untreated AN | 0.64 [0.17–1.63] | 9.52 | 5.36 | 0.067 (0.025–<br>0.182) | 0.119 (0.035–<br>0.407) | <0.001 |
| With untreated<br>AN | 4.58 [1.68–9.96] | 9.52 | 5.36 | 0.481 (0.211–<br>1.093) | 0.854 (0.287–<br>2.541) | 0.080 |
| B. Aneurysm-based analysis (comparison with SUAve) |  |  |  |  |  |  |
| Subgroup | Rate in This Study<br>(/1,000 PY, 95% CI, Byar) | SUAve Rate (/1,000<br>PY) |  | RR vs SUAve<br>(95% CI) | p-value |  |
| Overall | 1.18 (0.57–2.18) | 4.51 (1.81–9.29) |  | 0.263 (0.100–0.690) | 0.006 |  |
| No untreated AN | 0.58 (0.16–1.49) | 4.51 (1.81–9.29) |  | 0.130 (0.038–0.442) | 0.001 |  |
| With untreated AN | 3.88 (1.42–8.44) | 4.51 (1.81–9.29) |  | 0.861 (0.289–2.561) | 0.800 |  |
| C. Comparison with general population of Tochigi Prefecture |  |  |  |  |  |  |
| Outcome/Subgroup | Crude ratio (Observed/Expected, 95% CI) | Age-adjusted SIR (95% CI) |  | p-value |  |  |
| SAH — Overall | 9.74 (4.67–17.92) | 6.62 (3.17–12.17) |  | <0.001 |  |  |
| SAH — No untreated AN | 4.75 (1.29–12.15) | 3.22 (0.88–8.25) |  | 0.075 |  |  |
| All-stroke — Overall | 2.40 (1.70–3.30) | 2.11 (1.50–2.90) |  | <0.001 |  |  |

Incidence data for malignant neoplasms and cardiovascular diseases were obtained from the Ministry of Health, Labour and Welfare of Japan, and for dementia from the Hisayama Study. Age-adjusted SIRs were computed only for SAH and all-stroke using age-stratified incidence rates from Tochigi Prefecture; for all other endpoints, only crude (unadjusted) ORs versus the general population were calculated because age-specific population rates were unavailable. Rates are per 1,000 PY with 95% CIs by Byar's approximation; ORs were calculated using Wald methods.
Abbreviations: AN = aneurysm; RR = rate ratio; SIR = standardized incidence ratio; PY = person-years; CI = confidence interval.

Kaplan–Meier analysis demonstrated SAH-free survival rates of 99.4% at 5 years and 98.4% at 10 years in the overall cohort. In patients without untreated aneurysms, SAH-free survival rates were 99.6% at 5 years and 98.3% at 10 years, whereas in those with untreated aneurysms, rates were 97.0% and 94.9%, respectively. This difference was statistically significant on log-rank testing (p<0.05; Figure 1).

**Figure 1.**
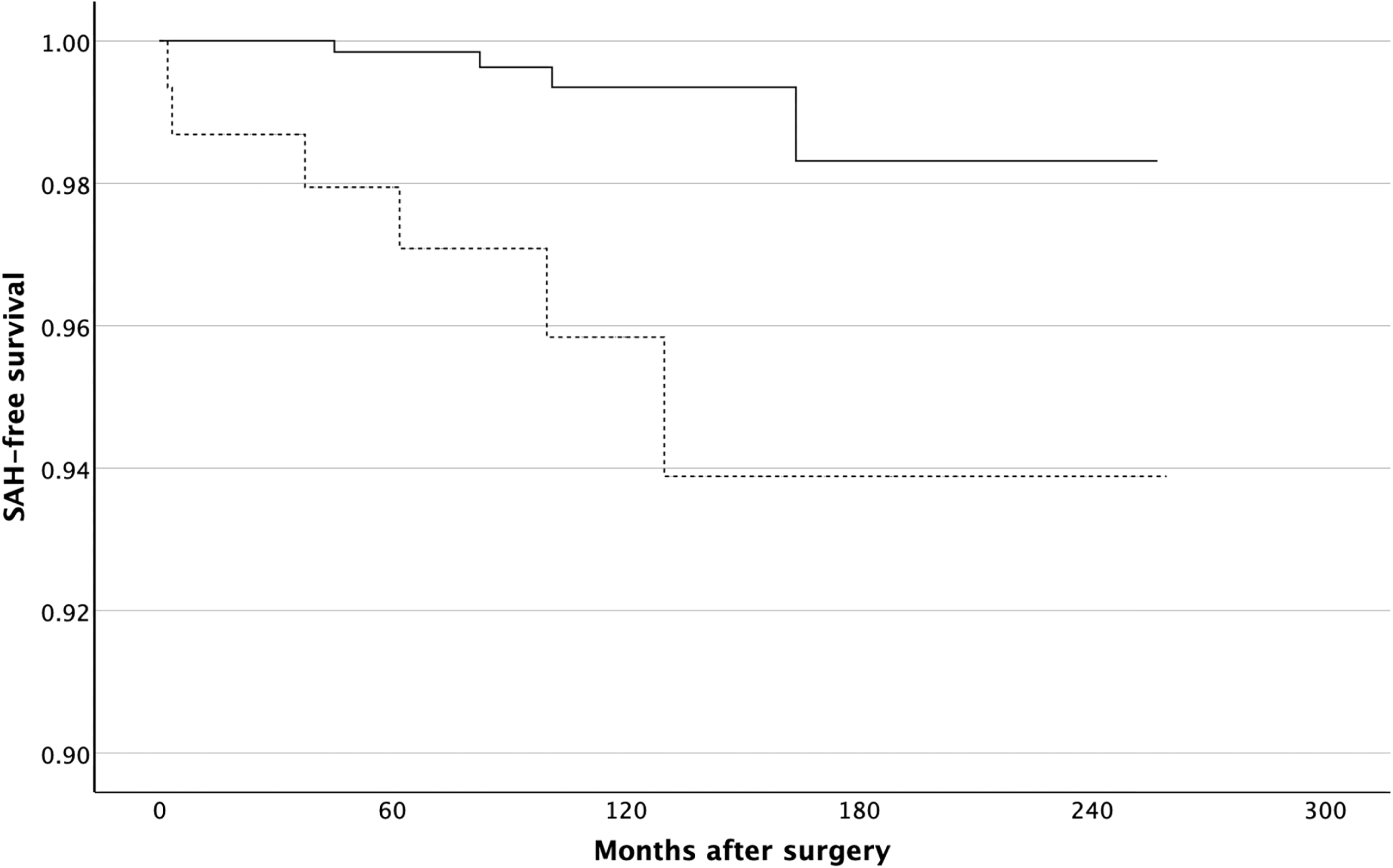
Kaplan–Meier curves of SAH-free survival stratified by the presence of untreated aneurysms. The solid line represents patients without untreated aneurysms (n = 782), and the dotted line represents patients with untreated aneurysms (n = 148). During follow-up, SAH-free survival was significantly lower in patients with untreated aneurysms (log-rank test, p < 0.05). The x-axis indicates months after surgery, and the y-axis indicates SAH-free survival probability.

**Figure 2.**
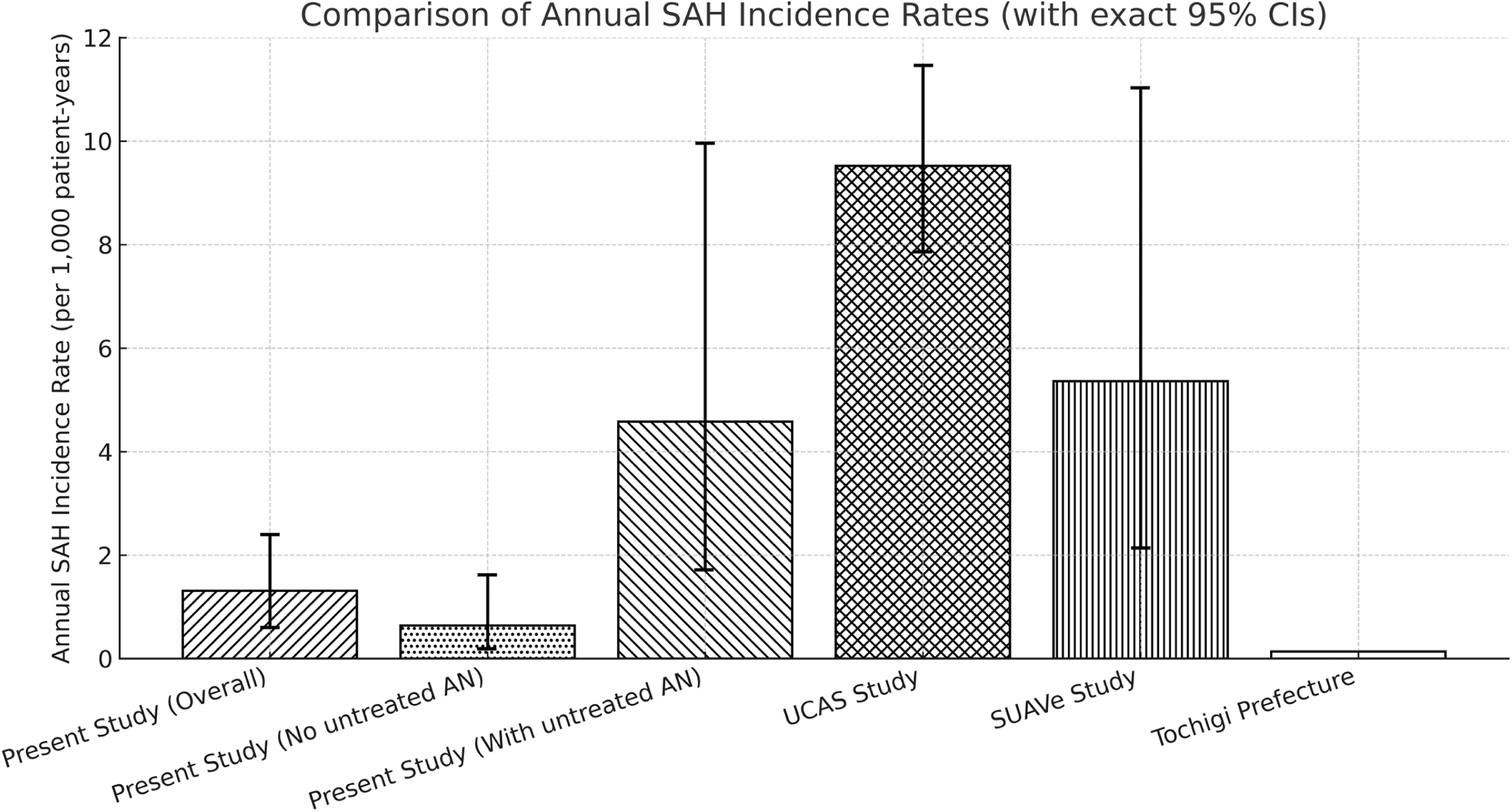
Comparison of Annual Incidence Rates of SAH. Annual SAH incidence rates are shown for six groups: the overall present study cohort (1.31/1000 PY, horizontal stripes), patients without untreated aneurysms (0.64/1000 PY, dotted), patients with untreated aneurysms (4.58/1000 PY, diagonal stripes), the UCAS study (9.52/1000 PY, solid black), the SUAVe study (5.36/1000 PY, vertical stripes), and the general population of Tochigi Prefecture (0.137/1000 PY, open bars). The vertical axis indicates annual SAH incidence rates (per 1000 patient-years), and error bars represent 95% confidence intervals (exact Poisson method).

### Subgroup and Risk Stratification Analyses

To further characterize heterogeneity of risk within the study cohort, subgroup analyses were performed. SAH incidence rates were lower in younger patients and in those with a single aneurysm, whereas older age and multiplicity were associated with increased long-term risk. Specifically, the incidence was 0.80 per 1,000 patient-years (95% CI, 0.20–2.10; Byar) in patients aged <60 years and 1.70 per 1,000 patient-years (95% CI, 0.90–3.20; Byar) in those aged ≥60 years. Similarly, incidence was 0.90 per 1,000 patient-years (95% CI, 0.40–1.80; Byar) in patients with a single aneurysm and 2.30 per 1,000 patient-years (95% CI, 1.10–4.50; Byar) in those with multiple aneurysms. These findings suggest that, even in this highly selected cohort with overall low rupture risk, patient-specific factors such as age and aneurysm multiplicity remain important determinants of long-term SAH risk.

### Comparison with Natural History Studies and General Population

Compared with the UCAS and SUAVe cohorts, the incidence of SAH in our series was substantially lower on both a patient- and aneurysm-based scale (Table 3). To partially account for age distribution differences across studies, we performed an indirect standardization using age-specific person-years from our cohort and the overall rupture rates reported in UCAS and SUAVe. This “pseudo–age-adjusted” comparison confirmed the robustness of our findings, indicating that the risk of SAH after clipping remained markedly reduced relative to natural history.

When compared with the general population of Tochigi Prefecture, however, the incidence of SAH in the overall cohort was significantly higher (SIR 6.62, 95% CI, 3.17–12.17; Byar; p < 0.001). Even when restricted to patients in the no untreated aneurysm group, the incidence remained elevated but did not reach statistical significance (SIR 3.22, 95% CI, 0.88–8.25; Byar; p = 0.075). Given the limited number of events, the confidence interval was wide and included the null value. The overall incidence of all-stroke was also significantly higher than that of the general population (SIR 2.11, 95% CI, 1.50–2.90; Byar; p < 0.001).

### Sensitivity Analyses

Bootstrap resampling confirmed the stability of incidence rate estimates, with confidence intervals similar to those obtained using Byar’s approximation. Bayesian analysis indicated a posterior probability exceeding 95% for a reduced risk of SAH compared with the UCAS cohort. Power analysis showed that the study had 80% power to detect a 50% reduction in SAH incidence relative to UCAS. For the borderline comparison with the general population (SIR = 3.22, 95% CI, 0.88–8.25; Byar; p = 0.075), the minimum detectable effect size was larger than the observed threefold increase, suggesting that although the difference did not reach statistical significance, the elevated risk remains clinically meaningful.

### Stroke Incidence

All-stroke events (SAH, cerebral infarction, and intracerebral hemorrhage) occurred in 38 cases, with an annual incidence rate of 4.98 per 1,000 patient-years (95% CI, 3.52– 6.83; Byar). The breakdown was 22 cases of cerebral infarction (2.88/1,000 patient-years; 95% CI, 1.81–4.36; Byar), 10 cases of SAH (1.31/1,000 patient-years; 95% CI, 0.63–2.41; Byar), and 6 cases of intracerebral hemorrhage (0.79/1,000 patient-years; 95% CI, 0.29–1.73; Byar). Compared with the general population of Tochigi Prefecture, the incidence of all-stroke was significantly higher without age adjustment (OR 2.41, 95% CI, 1.75–3.32; p<0.001) and remained significantly elevated after age adjustment (SIR 2.11, 95% CI, 1.50–2.90; Byar; p<0.001; Table 3).

### Other Disease Incidence and Mortality

Non-cerebrovascular disease incidence rates are summarized in Table 4. Malignant neoplasms occurred at 10.7 per 1,000 person-years (95% CI, 8.5–13.3), representing a significant excess compared with population references. Cardiovascular disease occurred at 2.1 per 1,000 person-years (1.2–3.4) with no significant difference versus references. Malignant and benign brain tumors were rare events and did not differ significantly from population rates. In contrast, dementia incidence in the ≥70-year subset was 11.0 per 1,000 person-years (6.5–17.1), significantly lower than in community-based cohorts.

**Table 4.** Annual Disease Incidence Rates (Comparison with General Population)

| Disease | Observed | Reference source | Observed rate<br>(/1,000py) | Reference rate<br>(/1,000py) | SIR /<br>RR | 95% CI<br>lower | 95% CI<br>upper | p-value |
| --- | --- | --- | --- | --- | --- | --- | --- | --- |
| Malignant<br>neoplasms | 81 | Cancer Statistics<br>Japan 2021 | 10.74 | 7.88 | 1.36 | 1.08 | 1.69 | 0.009 |
| Cardiovascular<br>disease | 16 | CIRCS | 2.12 | 1.5 | 1.41 | 0.81 | 2.27 | 0.221 |
| Malignant brain<br>tumors | 2 | Cancer Statistics<br>Japan 2021 | 0.27 | 0.046 | 5.76 | 0.65 | 20.14 | 0.096 |
| Benign brain<br>tumors | 4 | BTRJ | 0.53 | 0.15 | 3.54 | 0.95 | 8.78 | 0.056 |
| Dementia (≥70y<br>subset) | 18 | Hisayama Study | 10.96 | 32.3 | 0.34 | 0.2 | 0.53 | <0.001 |

Incidence rates for malignant neoplasms and malignant brain tumors were obtained from Cancer Statistics in Japan 2021<sup>20</sup>, for cardiovascular disease from CIRCS<sup>21</sup>, for dementia from the Hisayama Study<sup>22</sup>, and for benign brain tumors from BTRJ<sup>23</sup>. All reference rates were harmonized to per 1,000 person-years. Standardized incidence ratios (SIRs) or rate ratios (RRs) were calculated as observed events divided by expected events (expected = reference rate × person-years / 1,000). Ninety-five percent confidence intervals (CIs) were derived using Byar's approximation, and two-sided exact Poisson p-values were reported without multiplicity adjustment because all analyses were exploratory. Dementia analyses were restricted to the ≥70-year subset to align with Hisayama Study reporting (≥65 years).
Abbreviations: CI = confidence interval; SIR = standardized incidence ratio; RR = rate ratio.

During follow-up, 39 deaths occurred (5.2 per 1,000 person-years; 95% CI, 3.7–7.0), mainly from malignant neoplasms (30.8%), accidents (12.8%), and stroke (12.8%). Detailed standardized incidence ratios (SIRs), rate ratios (RRs), p-values, and age-band–specific analyses, as well as comparative statistics for non-SAH mortality versus the UCAS cohort (OR 0.429, 95% CI, 0.297–0.618, p<0.001), are provided in Table 4.

## DISCUSSION

### Effectiveness of Surgical Treatment

The main finding of this study is that direct surgery for anterior circulation UIAs significantly reduces SAH occurrence risk compared with natural history. In particular, in completely treated cases (group without untreated aneurysms), SAH occurrence risk was reduced by more than 93% compared with the UCAS study, demonstrating the effectiveness of prophylactic treatment. This result is consistent with reports by Molyneux et al.^25^ and Naggara et al.^26^, supporting the long-term effectiveness of direct surgery under appropriate case selection. Additionally, in a recent review Kim et al. emphasized that direct surgery is superior to endovascular treatment in terms of sustained long-term occlusion rates and low retreatment rates^27^, which is consistent with our study results, similar to these previous studies.

On the other hand, no significant difference from natural history was found in the patient group with untreated aneurysms, which was an anticipated result, as SAH incidence rates are likely to follow the natural history when untreated aneurysms remain.

### Persistent Cerebrovascular Risk

A noteworthy finding is that even in the patient group without untreated aneurysms, the incidence rates of cerebrovascular diseases including SAH were higher than in the general population. In the analysis of patients without untreated aneurysms, the incidence of SAH was significantly higher in the unadjusted analysis. However, after age adjustment, the standardized incidence ratio (SIR) was 3.22 (95% CI, 0.88–8.25; Byar; p = 0.075). Although the age-adjusted SIR exceeded 1 (3.22), the 95% CI included unity and the comparison did not meet conventional thresholds for statistical significance (p = 0.075). Accordingly, we interpret this as a suggestive, hypothesis-generating signal rather than definitive evidence. The upper bound (8.25) allows for a potentially important excess risk, whereas the lower bound (0.88) is compatible with little or no difference; more events and larger cohorts are required to refine this estimate. Sensitivity analyses were directionally consistent but did not alter this conclusion. In line with contemporary reporting guidance, we emphasize effect sizes with confidence intervals and avoid definitive claims based solely on proximity to arbitrary p-value thresholds.

The all-stroke incidence rate being about twice that of the general population likely reflects the high prevalence of vascular risk factors such as hypertension (61.4%) and smoking (19.7%). Rinkel et al.^28^ reported an increased cerebrovascular risk in families of SAH patients, suggesting genetic predisposition and structural vascular vulnerability. Our findings align with previous reports: Hokari et al. ^29^ observed increased cerebrovascular events after clipping, and Nieuwkamp et al.^30^ reported excess mortality and cardiovascular events in long-term SAH survivors. Early studies by Tsutsumi et al.^31^ and Matsumoto et al.^32^ demonstrated the preventive effect of surgery compared with conservative treatment, while Matsukawa et al.^33^ noted that some risk persists postoperatively. The annual SAH incidence rate of 1.31/1,000 patient-years in our cohort (95% CI, 0.63–2.41; Byar) is consistent with these reports.

Overall, surgical treatment markedly reduces SAH risk compared with natural history but does not normalize cerebrovascular risk to general population levels. These results underscore the importance of continued risk management and long-term surveillance after surgery.

### Non-cerebrovascular Disease Outcomes

Despite long-term follow-up exceeding 7,500 person-years, direct microsurgical clipping under general anesthesia did not lead to any clinically meaningful increase in the risk of major systemic diseases. Cardiovascular disease incidence was not elevated compared with representative Japanese population references, and dementia incidence in the ≥70-year subset was significantly lower than in community-based cohorts such as the Hisayama Study. These findings suggest that neither surgical intervention nor perioperative management adversely affects long-term systemic health, while ongoing medical follow-up may even contribute to earlier detection and management of cognitive decline.

The modest excess risk observed for malignant neoplasms (SIR 1.36) requires cautious interpretation. Patients who undergo clipping typically receive regular imaging, laboratory testing, and clinical visits for many years after surgery. This intensive medical surveillance increases the likelihood of diagnosing subclinical or early-stage cancers that would remain undetected in the general population. While this intensive surveillance likely explains most of the observed excess risk, we cannot entirely exclude the possibility that the more frequent use of CT imaging in this health-conscious group may have contributed minimally to the risk through cumulative radiation exposure. However, current epidemiological evidence suggests that the increased risk from diagnostic imaging doses is limited, particularly in adults. Thus, surveillance bias remains the more plausible explanation^34^. Indeed, no biological mechanism plausibly links a single neurosurgical intervention to de novo malignancy years later. Prior cohort studies have also attributed small cancer excesses to differences in diagnostic intensity rather than causality.

Taken together, our results demonstrate that direct microsurgical clipping effectively prevents SAH without imposing additional long-term systemic disease burden, and the apparent increase in cancer incidence most likely reflects differences in medical surveillance rather than true treatment-related risk.

### Comparison with Contemporary Literature

Our findings are broadly consistent with recent meta-analyses indicating that microsurgical clipping, when applied to appropriately selected patients, achieves durable aneurysm occlusion with very low retreatment rates. This reinforces the role of clipping as a definitive treatment strategy under optimal surgical conditions. However, direct comparison with endovascular series remains difficult, as most endovascular cohorts include older patients, posterior circulation aneurysms, or cases with higher procedural risk, and follow-up durations are often shorter. These differences in baseline risk and surveillance intensity limit strict comparability between treatment modalities. Importantly, the observed persistent elevation in cerebrovascular disease risk—approximately a twofold increase in stroke incidence compared with the general population—mirrors findings from previous long-term observational studies. This excess risk likely reflects the underlying vascular vulnerability of patients with UIAs rather than a direct consequence of clipping surgery itself. Clinically, this highlights the necessity of ongoing vascular risk factor management and structured follow-up, even after successful aneurysm treatment.

## LIMITATIONS

This study has several important limitations. First, its single-center, retrospective design and highly selective inclusion criteria—restricted to anterior circulation UIAs judged to carry minimal operative risk and excluding giant aneurysms—represent a best-case surgical scenario. Therefore, the findings may not be generalizable to broader UIA populations, including patients with posterior circulation or complex aneurysms or institutions with different surgical expertise or treatment philosophies.

Second, comparisons with natural history cohorts such as UCAS and SUAVe should be interpreted cautiously because those studies included all detected UIAs regardless of treatability, whereas our surgical cohort comprised a highly selected subset with intrinsically lower baseline risk, introducing inevitable differences in baseline comparability.

Third, although long-term imaging follow-up was achieved in approximately 90% of patients at both 5 and 10 years, some attrition occurred over time. Importantly, not all censored cases represented true loss to follow-up; many were discontinued for documented reasons such as advanced age, intercurrent illness, or relocation. Approximately 100 patients were true lost-to-follow-up cases, which could have introduced bias, particularly for late complications. However, sensitivity analyses assuming worst-case scenarios—in which all missing patients experienced the event—increased the estimated recurrence rate but did not fundamentally alter the temporal pattern, supporting the robustness of our conclusions.

Fourth, certain outcomes—particularly mild cognitive changes, psychosocial impacts, or subtle quality-of-life impairments—were not systematically assessed during routine clinical follow-up and may therefore have been underestimated.

Fifth, the limited number of recurrence and SAH events constrained statistical power for subgroup analyses, especially location-specific comparisons, and precluded meaningful multivariable modeling; thus, observed differences should be considered hypothesis-generating rather than definitive.

Finally, nationwide age-specific incidence tables contemporaneous with our follow-up were limited or unavailable for several endpoints. We therefore used representative Japanese registries (Cancer Statistics in Japan, CIRCS, Hisayama, BTRJ) as external references, harmonizing all rates to per 1,000 person-years. Differences in endpoint definitions (e.g., our cardiovascular disease endpoint includes ischemic heart disease plus heart-failure hospitalizations, whereas CIRCS reports ischemic heart disease only), age structures (e.g., Hisayama reports ≥65y dementia incidence; we used the ≥70y subset), and calendar periods may have introduced non-differential misclassification and limited direct comparability. Accordingly, all population comparisons are presented with 95% CIs and exact p-values in Table 4, emphasizing effect sizes over strict hypothesis testing.

### Clinical Implications and Decision-Making Framework

The results of this study provide several important clinical implications. First, even small aneurysms may warrant consideration for preventive treatment, particularly in younger patients, in whom the potential long-term benefits could outweigh the procedural risks. Second, continuous cerebrovascular risk management is necessary even after surgical aneurysm treatment. Strict control of vascular risk factors such as hypertension, dyslipidemia, and diabetes is important to mitigate the residual risk of cerebrovascular disease. Third, long-term imaging surveillance is essential to detect de novo aneurysm formation and growth of untreated aneurysms. The fact that SAH from de novo aneurysms accounted for 40.0% of events in this study supports the need for continued vigilance.

Furthermore, while cerebrovascular disease risk remains elevated, other disease risks such as malignant neoplasms and cardiovascular disease were comparable to the general population. Therefore, balanced and comprehensive health management is warranted. Overtreatment should be avoided, but maintaining a system for early detection and treatment through regular health checks is likely to lead to optimal outcomes. Based on these findings, a pragmatic framework for clinical decision-making in asymptomatic UIAs can be proposed:

- **High-benefit candidates**: Young patients (<70 years) with anterior circulation aneurysms that are small, have favorable neck morphology, and are surgically accessible may derive the greatest long-term benefit from prophylactic clipping.
- **Risk–benefit equilibrium**: Middle-aged patients (70–80 years) with otherwise favorable anatomy require individualized assessment. In this group, decisions should weigh surgical durability against comorbidities, vascular risk profiles, and life expectancy.
- **Conservative management**: Elderly patients (>80 years) or those with complex aneurysm morphology, posterior circulation aneurysms, or high operative risk may be better served by conservative observation or, when technically feasible, endovascular treatment.

It should be emphasized that these recommendations reflect outcomes under our highly selective surgical criteria and may not be directly applicable to all institutions or patient populations. Nonetheless, this framework underscores the need to balance rupture risk reduction with procedural safety, durability, and patient-centered values when determining optimal management for UIAs. These points should be considered when individualizing treatment strategies for asymptomatic UIAs.

### Comparative Analysis

Taken together, our comparative analysis highlights two contrasting perspectives. First, when benchmarked against natural history cohorts (UCAS and SUAVe), the postoperative incidence of SAH was dramatically lower, even after indirect age standardization. This finding underscores the long-term protective effect of clipping in carefully selected patients.

Second, however, when compared with the general population of Tochigi Prefecture, the incidence of SAH in our operated cohort remained significantly higher (SIR 6.62, p<0.001). Importantly, this excess risk persisted—albeit at borderline significance—when limited to patients without untreated aneurysms (SIR 3.22, p=0.075). The overall incidence of stroke was also significantly higher than that of the general population (SIR 2.11, p<0.001). These results suggest that while microsurgical clipping reduces the risk relative to untreated natural history, the operated population continues to carry a residual vulnerability to SAH and stroke, possibly reflecting the development of de novo aneurysms or concomitant vascular risk factors.

## CONCLUSIONS

Microsurgical clipping for anterior circulation UIAs demonstrates significant effectiveness in reducing SAH occurrence risk compared to natural history, with a 93% risk reduction achieved in patients without untreated aneurysms. However, this study’s highly selective patient criteria—including only cases judged to carry minimal operative risk—critically limits the generalizability of these favorable outcomes to broader UIA populations.

Several key findings warrant emphasis: First, even after successful surgical treatment, patients maintained approximately 2-fold elevated cerebrovascular disease risk compared to the general population, indicating persistent underlying vascular vulnerability that requires ongoing management. Second, de novo aneurysms accounted for 40% of post-surgical SAH events, emphasizing that the vascular substrate for aneurysm formation persists despite successful treatment of index lesions. Third, non-cerebrovascular outcomes remained comparable to the general population, demonstrating that microsurgical intervention does not compromise overall health status when performed under optimal conditions.

The clinical implications are nuanced: while prophylactic surgery can be highly effective under optimal conditions, treatment decisions must carefully weigh patient-specific factors, institutional expertise, and alternative management strategies. The persistent vascular risk necessitates comprehensive long-term management including rigorous control of modifiable risk factors such as hypertension and smoking. These findings support microsurgical treatment as an effective option for appropriately selected anterior circulation UIAs while emphasizing that patient selection criteria, institutional capabilities, and comprehensive post-treatment care are critical determinants of long-term success. Future research should focus on developing refined risk stratification models and conducting comparative effectiveness studies across different treatment modalities in similar patient populations.

## Data Availability

All data supporting the findings of this study are available within the article and its supplementary materials. De-identified individual participant data are available from the corresponding author upon reasonable request.

## Acknowledgments

The authors thank the neurosurgical team and radiology staff for their contributions to patient care and data collection.

